# The association between SARS-CoV-2 infection and neuronal damage: A population-based nested case-control study

**DOI:** 10.1101/2021.09.02.21263019

**Authors:** N. Ahmad Aziz, Marina L.S. Santos, Monique M.B. Breteler

## Abstract

**Objective:** To assess whether severe acute respiratory syndrome coronavirus-2 (SARS-CoV-2) infection is associated with changes in plasma levels of neurofilament light chain (NfL), an extremely sensitive marker of neuroaxonal damage, in community-dwelling individuals.

**Setting:** This study was embedded within the Rhineland Study, an ongoing community-based cohort study in Bonn, Germany

**Design:** Cross-sectional nested case-control study.

**Participants:** Participants were selected based on results from a previously conducted seroprevalence survey within the framework of the Rhineland Study. Cases were defined as those individuals who had had two positive confirmatory test results, including a recombinant spike-based immunofluorescence assay and a plaque reduction neutralization test (N=21). As controls, a random sample of individuals with a negative ELISA test result (Controls I, N=1117), and those with a borderline or positive ELISA test result who failed confirmatory testing (Controls II, N=68), were selected.

**Outcome measures:** Plasma levels of NfL at the time of measurement, as well as change in plasma NfL levels compared to previously measured pre-pandemic levels

**Results:** After adjustment for age, sex and batch effects, serologically confirmed SARS-CoV-2 infection was neither associated with cross-sectional NfL levels, nor with the magnitude of change from pre-pandemic levels, compared to either of the two control groups. Similarly, after adjustment for age, sex and batch effects, self-reported neurological symptoms – including altered sense of smell or taste, headache, myalgia and fever – were not associated with changes in NfL levels in participants with a serologically confirmed SARS-CoV-2 infection (all p ≥ 0.56).

**Conclusions:** Our findings indicate that mild-to-moderate coronavirus disease-19 is unlikely to be associated with a clinically relevant degree of neuroaxonal damage, even in those cases associated with neurological symptoms.

## INTRODUCTION

Neurological signs and symptoms are reported in more than one-third of coronavirus disease-19 (COVID-19) patients, sometimes even as the presenting or sole manifestation of the disease.^1, 2^ Although (para-)infectious neuronal injury has been demonstrated in hospitalized cases with severe to critical illness,^3-9^ it is still unclear whether it also occurs in patients with mild-to-moderate disease in whom neurological symptoms are also very common.^1^ Therefore, we assessed whether severe acute respiratory syndrome coronavirus-2 (SARS-CoV-2) infection is associated with changes in plasma levels of neurofilament light chain (NfL), an ultrasensitive marker of neuroaxonal damage, in community-dwelling individuals.

## METHODS

Between April 24^th^ and June 30^th^, 2020, we conducted a seroprevalence survey embedded within the Rhineland Study, an ongoing community-based cohort study in Bonn, Germany, the results of which have been published previously.^10^ SARS-CoV-2 antibodies were measured with an enzyme-linked immunosorbent assay (ELISA), followed by confirmatory testing of borderline and positive test results with a recombinant spike-based immunofluorescence assay and a plaque reduction neutralisation test.^10^ We defined cases as those participants who had two positive confirmatory test results (N=21). As controls we selected a random sample of individuals with a negative ELISA (Controls I, N=1117), and those with a borderline or positive ELISA who failed confirmatory testing (Controls II, N=68). Plasma NfL levels were measured with the Quanterix Simoa NF-light® assay in samples collected during the serosurvey, as well as in samples from the same individuals collected between 2016 and 2019. Generalized linear (mixed-effect) models were used to assess the association between SARS-CoV-2 infection and log-transformed NfL levels, both cross-sectionally and as change from pre-pandemic levels.

## RESULTS

Participant characteristics are displayed in **Table 1**. After adjustment for age, sex and batch effects, serologically confirmed SARS-CoV-2 infection was neither associated with cross-sectional NfL levels, nor with the magnitude of change from pre-pandemic levels, compared to either of the two control groups (**Figure 1**). Similarly, after adjustment for age, sex and batch effects, self-reported neurological symptoms – including altered sense of smell or taste, headache, myalgia and fever – were not associated with changes in NfL levels in participants with a serologically confirmed SARS-CoV-2 infection (all p ≥ 0.56). Sensitivity analysis by excluding NfL levels above 4 standard deviations from the control groups did not materially change any of the results (data now shown).

**Table 1.** Characteristics of the study population.

|  |  | *SARS-CoV-2 infection*<sup>*</sup> | *Control group I* | *Control group II* | *P-value*<sup>**</sup> |
| --- | --- | --- | --- | --- | --- |
| N |  | 21 | 1117 | 68 |  |
| Age [years] (SD) |  | 52.3 (13.5) | 53.9 (13.7) | 57.5 (14.6) | 0.220 |
| Sex (% female) |  | 67 | 57 | 44 | 0.070 |
| Body weight [kg] (SD) |  | 70.9 (11.4) | 76.0 (15.9) | 77.7 (15.9) | 0.220 |
| Mean number of symptoms (SD) |  | 9.5 (5.9) | 5.7 (4.8) | 5.7 (4.8) | 0.002 |
| Olfactory dysfunction (%) |  | 33 | 6 | 7 | <0.001 |
| Gustatory dysfunction (%) |  | 38 | 5 | 9 | <0.001 |
| Headache (%) |  | 43 | 44 | 47 | 1.00 |
| Myalgia (%) |  | 48 | 28 | 28 | 0.200 |
| Fever in last month (%) |  | 10 | 2 | 1 | 0.030 |
| Fever before last month (%) |  | 29 | 9 | 6 | 0.009 |
| Plasma NfL [pg/mL]<sup>***</sup> | Mean (SD) | 9.3 (4.3) | 10.8 (7.0) | 9.9 (7.3) | 0.610 |
|  | Median (IQR) | 7.9 (6.7 – 11.5) | 8.3 (6.7 – 12.3) | 8.0 (5.8 – 11.6) |  |
|  | Range | 3.9 – 22.3 | 3.9 – 52.3 | 2.2 – 91.9 |  |

\*) In our serosurvey we identified 22 individuals with serological confirmation of a sustained SARS-CoV-2 infection; however, in one of these individuals there was an insufficient amount of plasma left to allow measurement of NfL levels.
\*\*) Based on an analysis of variance or $\chi^2$ -test for continuous and categorical variables, respectively, except for plasma NfL levels. The p-value for plasma NfL is based on a linear regression model with log-transformed plasma NfL level as the outcome and serogroup as predictor, while adjusting for age, sex and batch effects (p-value for contrast with Control group I).
Abbreviations: IQR = interquartile range (i.e. the 25<sup>th</sup> and 75<sup>th</sup> percentiles) of the NfL distribution, N = number of individuals in each group, NfL = neurofilament light chain, SD = standard deviation.

**Figure 1.**
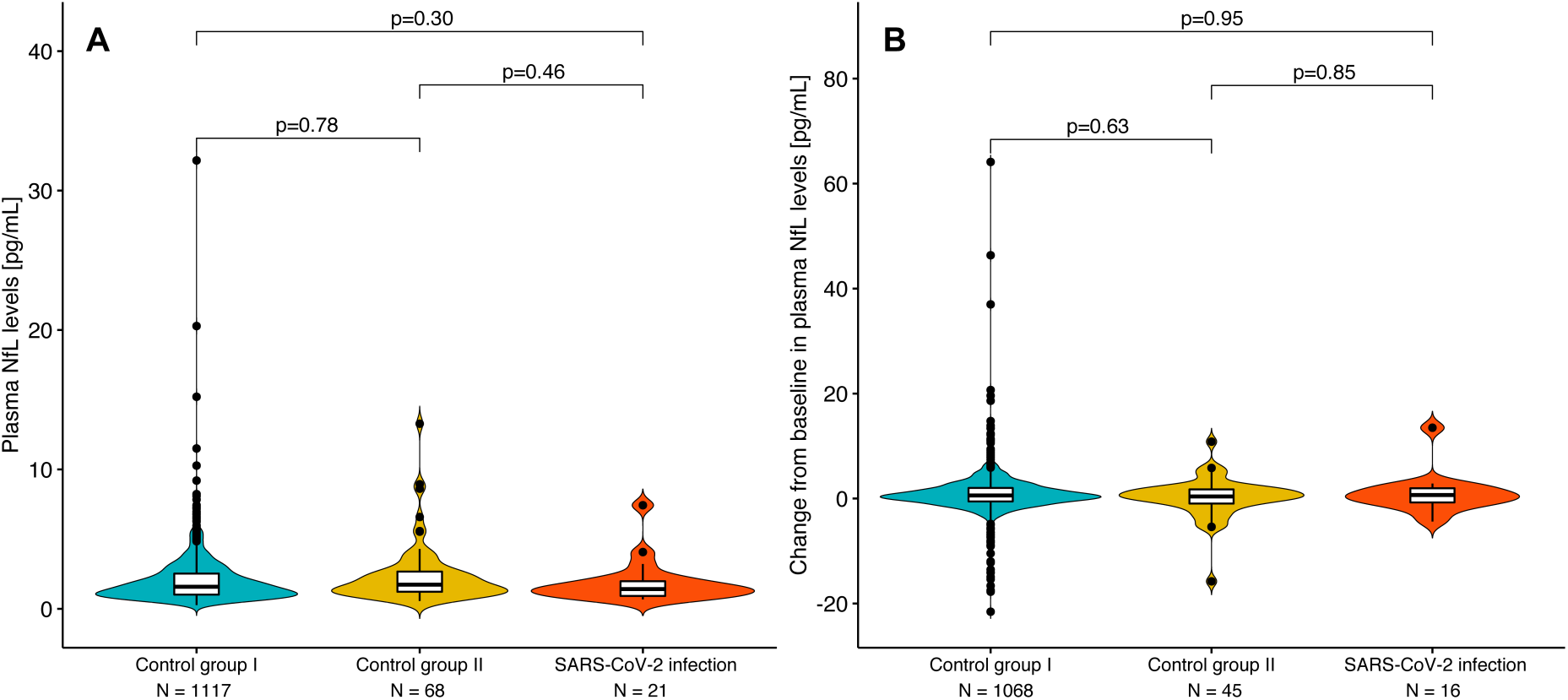
The association between SARS-CoV-2 infection and plasma neurofilament light chain (NfL) levels. Cross-sectional levels of plasma NfL did not differ between cases with SARS-CoV-2 infection and either of the two control groups, after adjustment for age, sex and batch effects (**A**). Similarly, after additional adjustment for the time passed between the two measurement occasions, intra-individual changes in plasma NfL did not differ between cases with SARS-CoV-2 infection and either of the two control groups (**B**). The violin-plots reflect the frequency distribution of NfL values, while the box-plots indicate the median (bold horizontal lines) and interquartile ranges; values outside 1.5 times the interquartile range from the median are represented by black dots. The p-values for cross-sectional and longitudinal intergroup comparisons were based on results from generalized linear and generalized linear mixed-effects models, respectively. The numbers below the horizontal axes represent the number of individuals included in each category; please note that in a small subset of individuals longitudinal measurements were not possible due to insufficient amounts of plasma.

## DISCUSSION

We present the first population-based study of the association between SARS-CoV-2 infection and neuronal injury. By implementing a serosurvey with a rigorous multi-tiered testing approach in an ongoing large prospective population-based cohort study, we were able to minimize the risk of false-positives and take advantage of existing bio-samples collected before the start of the pandemic.^10^ In community-dwelling individuals with mild-to-moderate symptoms, we found no association between serologically confirmed recent infections by SARS-CoV-2 and neuroaxonal damage, neither in comparison to individuals without evidence of a recent infection, nor in comparison to individuals with probable cross-reactive antibodies against other viruses.

Our results parallel those from a recent cross-sectional study in 2652 children, of whom 148 had serological evidene of a SARS-CoV-2 infection, which also did not find changes in blood NfL levels in pediatric cases with asysmptomatic to moderate COVID-19.^11^ However, these and our results contrast with those of a previous small-scale study that reported increased serum NfL levels in health care workers with mild-to-moderate COVID-19.^12^ This discrepancy may be accounted for by the fact that this other study did not assess intra-individual changes in NfL levels, only included a small reference group, and the results may have been confounded by incomplete information on participants’ age.^12^ Reports of neuroaxonal damage in hospitalized patients with severe COVID-19 may thus reflect neuronal injury due to dyshomeostasis and hypercoagulation engendered by the systemic infection and inflammation without direct viral neuroinvasion.^3-9^

Potential limitations of our study include the relatively small number of cases with a serologically confirmed SARS-CoV-2 infection and lack of data on the exact timing of the infection in seropositive individuals. However, given that the samples were collected shortly after the start of the pandemic in Germany, and the relatively long half-life of NfL that is estimated to be up to a few weeks,^13^ it is unlikely that we would have missed any substantial or progressive post-infectious elevations of serum NfL levels.

Our findings indicate that mild-to-moderate COVID-19 is unlikely to be associated with a clinically relevant degree of neuroaxonal damage, even in those cases associated with neurological symptoms like olfactory and gustatory dysfunction, headache and myalgia.

## Data Availability

The Rhineland Study's dataset is not publicly available because of data protection regulations. Access to data can be provided to scientists in accordance with the Rhineland Study's Data Use and Access Policy. Requests for further information or to access the Rhineland Study's dataset should be directed to.

## ACKNOWLEDGEMENTS

The authors would like to thank all the staff members and participants of the Rhineland Study, as well as our collaborators at the National Consultant Laboratory for Coronaviruses, especially Dr. Victor M. Corman, Dr. Marcel A. Müller and Prof. Dr. Christian Drosten, who conducted and supervised the SARS-CoV-2 testing procedures.

## CONTRIBUTORSHIP STATEMENT

N.A.A. and M.M.B.B. conceived and designed the study. N.A.A. and M.L.S. coordinated the data and bio-sample collection and processing. N.A.A. did the data analyses. N.A.A. and M.M.B.B. drafted the first version of the manuscript. All authors contributed to data interpretation, participated in revising the manuscript for important intellectual content, and read and approved the final manuscript.

## COMETING INTEREST

The authors report no conflicts of interest.

## ETHICS STATEMENT

Approval to undertake the Rhineland Study was obtained from the ethics committee of the University of Bonn, Medical Faculty (reference ID: 338/15).

